# ADVISE: A Machine Learning Framework for Early Recognition of a Surrogate Marker for Ventilator-Associated Pneumonia Using Routinely Collected Critical Care Data

**DOI:** 10.64898/2026.06.15.26355691

**Authors:** Nabeel Amiruddin, Sophie Mellor, Rehman Crisp, Ananya Nair, Maya Patel

**Affiliations:** Critical Care Unit, Russell’s Hall Hospital, The Dudley Group NHS Foundation Trust, Dudley, UK, DY1 2HQ

**Keywords:** Ventilator-associated pneumonia, machine learning, XGBoost, critical care, procalcitonin, ADVISE, NHS

## Abstract

**Background:** Ventilator-associated pneumonia (VAP) is the most frequent nosocomial infection in critical care, affecting 20-36% of mechanically ventilated patients. Early prediction is hampered by the absence of a reliable, objective diagnostic standard. We developed ADVISE (Automated Dudley Ventilation Infection Series Evaluation), a machine learning model to predict physiological deterioration consistent with developing VAP using routinely collected electronic health record data from a UK NHS intensive care unit.

**Methods:** Retrospective observational study of admissions at Russell’s Hall Hospital ICU (2008-2026). Following National Data Opt-Out exclusion (158 admissions, 4.2%), 3,566 admissions generated 33,208 candidate 48-hour observation blocks. Six temporal variables - FiO₂, ventilator mode, P:F ratio, procalcitonin (PCT), secretion amount, and secretion description - were extracted across the baseline window (hours 1-24). A composite VAP-surrogate outcome required concurrent P:F ratio decline (≥5%) and PCT rise (≥0.5 ng/mL) across the outcome window (hours 25-48). After sequential quality filters, 2,134 blocks (18 positive, 0.84% prevalence) were retained. An XGBoost classifier was trained using nested 5-fold cross-validation with scale_pos_weight=114.0 and ROC-based hyperparameter optimisation on 1,495 training blocks, evaluated on 639 held-out test blocks. Performance was assessed via AUROC, AUPRC, and calibration (Brier score). Bootstrap resampling (1,000 iterations) generated 95% confidence intervals.

**Results:** On the held-out test set (n=639, 5 positive outcomes), ADVISE achieved AUROC 0.874 [95% CI: 0.771-0.939] and AUPRC 0.031 [0.008-0.069], representing a 4.0-fold improvement over the no-skill baseline. Nested cross-validation mean AUROC was 0.844 ± 0.078 (range 0.716-0.915). At the Youden-optimal threshold, sensitivity was 0% with specificity 97.8%, reflecting extreme class imbalance (0.78% test prevalence). A threshold targeting 80% sensitivity achieved sensitivity 80.0% [33.3-100.0%], specificity 87.4% [84.8-89.9%], positive predictive value 4.8% [1.1-9.9%], and negative predictive value 99.8% [99.4-100.0%], detecting 4 of 5 VAP cases with approximately 80 false alarms (12.6% false positive rate). Brier score was 0.0078. Feature importance identified baseline P:F ratio as the dominant predictor (41.3% total gain), followed by ventilator mode (26.1%), secretion amount (13.2%), secretion description (9.1%), procalcitonin (5.9%), and FiO₂ (4.5%).

**Conclusions:** ADVISE demonstrates that baseline oxygenation trajectory and ventilatory support patterns - derived exclusively from routinely charted ICCA variables - can identify admissions at risk of VAP-related physiological deterioration with meaningful discrimination (AUROC 0.874) despite severe class imbalance. The 80% sensitivity operating point offers a clinically actionable alert rate (12.6% FPR), supporting integration into existing ICU workflows. This proof-of-concept study establishes feasibility; multi-site prospective validation is required before clinical deployment.

## Introduction

### Ventilator-Associated Pneumonia: Epidemiology and Clinical Burden

Ventilator-associated pneumonia (VAP) is defined as pneumonia arising 48 or more hours after endotracheal intubation and commencement of mechanical ventilation^3,7^. It is the most frequent nosocomial infection in critical care, affecting 20–36% of mechanically ventilated patients^1,2^ with a reported incidence of 2–16 episodes per 1,000 ventilator days^1,3^. VAP is associated with substantial morbidity and an attributable mortality of approximately 10% of all ITU patients^4^. Each episode costs the NHS an additional £6,000–£22,000 and increases ICU length of stay by approximately 28%^5^. The Dudley population served by Russell’s Hall Hospital includes areas of significant socioeconomic deprivation (Index of Multiple Deprivation deciles 1–3 comprising approximately 35% of the local population), which may influence generalisability. Despite national prevention initiatives, including the Intensive Care Society (ICS) Recommended Bundle^6^, VAP continues to impose significant cost and clinical burden on UK critical care services.

### Diagnostic Challenges

Accurate and timely diagnosis of VAP has proved persistently challenging. No universally accepted gold standard currently exists^7^. It is suggested that more than 50% of patients clinically diagnosed with VAP may not have true pneumonia, while up to one-third of confirmed cases may go undetected^8^. The Clinical Pulmonary Infection Score (CPIS) incorporating body temperature, peripheral leukocyte count, tracheal secretion character, oxygenation ratio (PaOL/FiOL), chest radiographic findings, and semi-quantitative microbiological culture results on a 0–12 scale^9,10^ provides a quantifiable bedside threshold (CPIS ≥6 indicating high probability of VAP), yet its reliability is limited by modest sensitivity and specificity and considerable inter-observer variability^10^.

### Prior Machine Learning Approaches

Early machine learning models for VAP prediction predominantly used conventional supervised learning on structured electronic health record data^12,13^, with many studies leveraging the publicly available MIMIC-III database^12–15^. Most treated patient data as static snapshots rather than longitudinal trajectories, limiting clinical applicability^13,14^. The PREDICT model applied long short-term memory (LSTM) networks to ICU time-series data, demonstrating the importance of capturing temporal physiological patterns^12^. However, a unifying limitation remains the absence of prospective, multi-site validation within NHS infrastructure^13,14^.

### Study Aims and Intended Clinical Use

ADVISE is designed for use by ICU clinicians and nurses as a passive background monitoring tool operating within the ICCA electronic patient record. It is intended to generate a daily risk flag for each mechanically ventilated patient, based on the preceding 24-hour physiological trajectory, to prompt clinical review. The model is not intended to replace clinical judgement or diagnostic workup, but to serve as an objective early-warning signal that can direct clinical attention toward higher-risk patients earlier in the course of deterioration. Prior to data extraction, the National Data Opt-Out register was consulted, and admissions of patients with active Type 1 opt-outs were excluded from the cohort (n=158 admissions, 4.2% of the 3,724 eligible), resulting in a final pre-screened cohort of 3,566 admissions.

This study aimed to: (1) implement a methodology for training a machine learning model to predict physiological deterioration consistent with VAP using only routinely charted variables from a UK NHS electronic patient record; (2) evaluate model discrimination and calibration on a held-out test set; (3) characterise the relative contribution of individual predictors; and (4) provide proof-of-concept for an NHS-deployable early-warning system.

## Method

### Study Design and Setting

This retrospective study analysed electronic health record data from Russell’s Hall Hospital Critical Care Unit, The Dudley Group NHS Foundation Trust, Dudley, UK (March 2008–February 2026). The 23-bed mixed medical-surgical ICU serves approximately 320,000 population in Dudley Metropolitan Borough. The study received Trust R&D approval (HRA REC: 26/HRA/0828) and Caldicott Guardian agreement, conducted per Declaration of Helsinki and UK Policy Framework for Health and Social Care Research.

### Inclusion and Exclusion Criteria

Patients receiving invasive mechanical ventilation^3,7^ ≥48 consecutive hours with continuous hourly documentation were included. Blocks were excluded if: (1) Procalcitonin(PCT) not measured in both baseline (hours 1–24) and outcome (hours 25–48) periods; (2) PCT >15 ng/mL at hour 24; or (3) median baseline PaO_2_:FiO_2_(P:F) ratio <13 kPa. Antibiotic therapy, sedation, neuromuscular blockade, prone positioning, and vasopressor use were not extracted (unmeasured confounding acknowledged in Limitations).

### Predictors and Data Source

Six predictors: FiOL, PCT, P:F ratio, secretion amount/description and ventilator mode were selected based on clinical knowledge, literature review, and routine ICCA availability. All data extracted from Philips ICCA. Variables: FiOL (0.21–1.00, hourly); PCT (ng/mL, daily); P:F ratio (kPa, automated); secretion characteristics (hourly, nurse-charted); ventilator mode (hourly). Chart times rounded to nearest hour. Only pseudonymised identifiers used.

### Data Processing

Physiologically implausible values flagged and replaced (<0.5% observations; Supplementary Methods). Categorical variables encoded on ordinal 0–1.0 scales (Supplementary Methods: Table S1). Missing data managed using LOCF/backward fill (stepwise variables) and linear interpolation (continuous variables). Residual missing values (<1%) replaced with training set medians (Supplementary Methods: Table S2). Continuous predictors not standardised (XGBoost invariant to monotonic transformations).

### Outcome Definition

Timelines divided into 48-hour blocks: baseline (hours 1–24) and outcome (hours 25–48), providing 24-hour prediction horizon. Primary outcome: composite VAP-surrogate requiring both (1) P:F ratio decline ≥5% from baseline to outcome AND (2) PCT increase ≥0.5 ng/mL from hour 24 to 48, capturing concurrent oxygenation and inflammation worsening. Outcome derived algorithmically without subjective interpretation.

### Model Development

XGBoost selected for: empirical performance on imbalanced data; computational efficiency; interpretability; no GPU requirement. Data stratified by outcome, split 70:30 (training/test). All 144 temporal features (6 variables × 24 timepoints) used.

Nested 5-fold cross-validation separated hyperparameter optimisation (inner) from performance estimation (outer). Class imbalance handled via scale_pos_weight=114.0 (114-fold weighting of positive cases). ROC metric used for optimization (robust to severe imbalance). Hyperparameter grid explored 192 combinations (Supplementary Methods: Table S3). Final hyperparameters: nrounds=200, max_depth=3, η=0.05, γ=1, colsample_bytree=0.8, min_child_weight=3, subsample=0.9. Final model trained on all 1,495 training observations. Classification threshold determined via Youden’s J statistic on training data.

### Statistical Analysis

Test set performance: AUROC, AUPRC, sensitivity, specificity, PPV, NPV, accuracy; Brier score; gain-based feature importance. Bootstrap resampling (1,000 iterations, stratified) generated 95% CIs. Calibration assessed via Cox method. Decision curve analysis not performed (insufficient positives, n=5). Analyses in R 4.1.0 (software: Supplementary Methods). Reported per TRIPOD+AI guidelines^18^ (Supplementary Table S1: compliance checklist).

**Figure 1.**
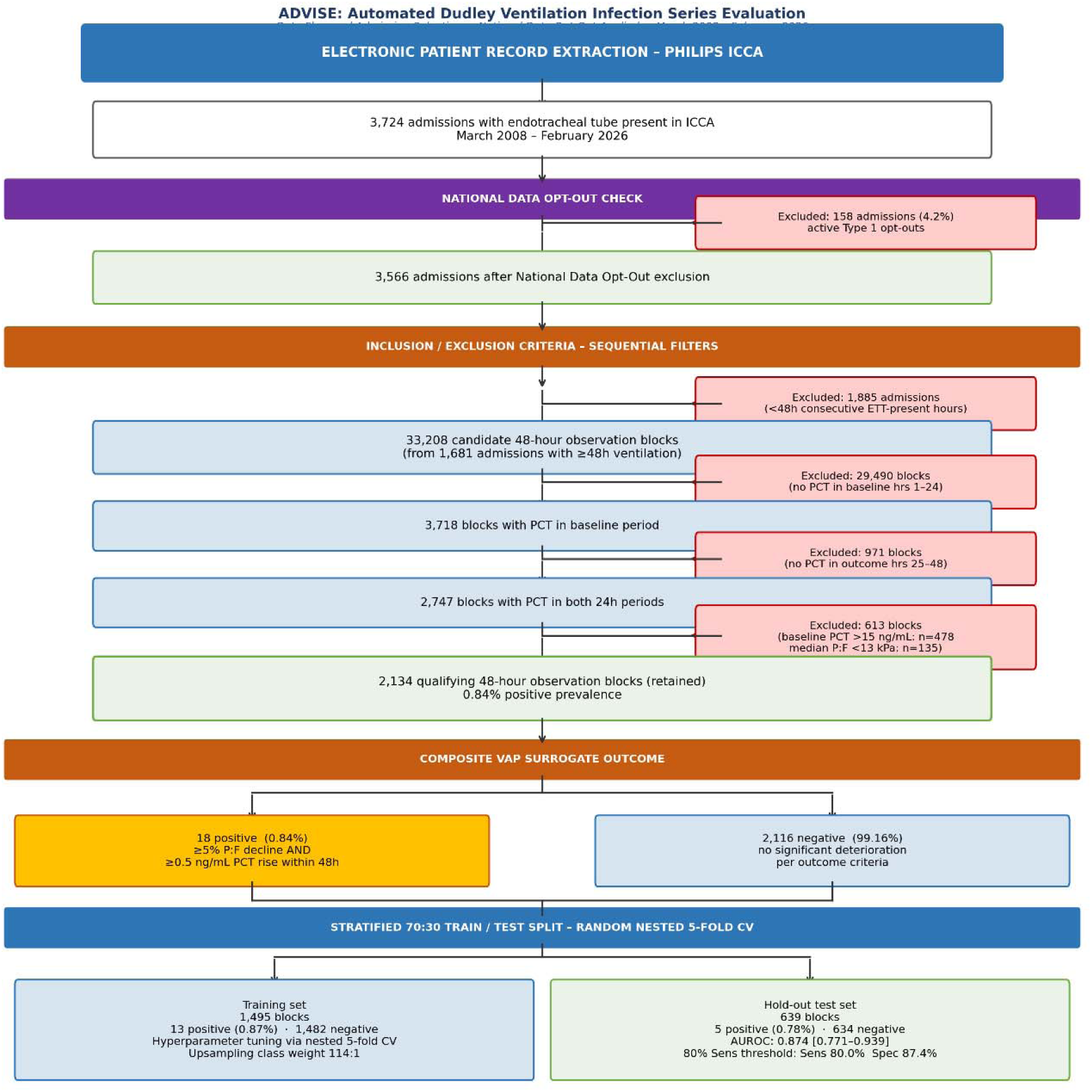
Patient flow diagram showing inclusion/exclusion criteria and final dataset composition.

## Results

Following National Data Opt-Out exclusion, 3,566 admissions (2008–2026) generated 33,208 candidate 48-hour blocks. Sequential filtering for procalcitonin availability in both the baseline and outcome periods (3,718 then 2,747 blocks), baseline PCT ≤15 ng/mL (2,269 blocks), and median baseline P:F ratio ≥13 kPa retained 2,134 observation blocks (18 positive outcomes, 0.84% prevalence). Blocks were split 70:30 into training (n=1,495; 13 positive) and test (n=639; 5 positive) sets, with all positive outcomes occurring post-2020.

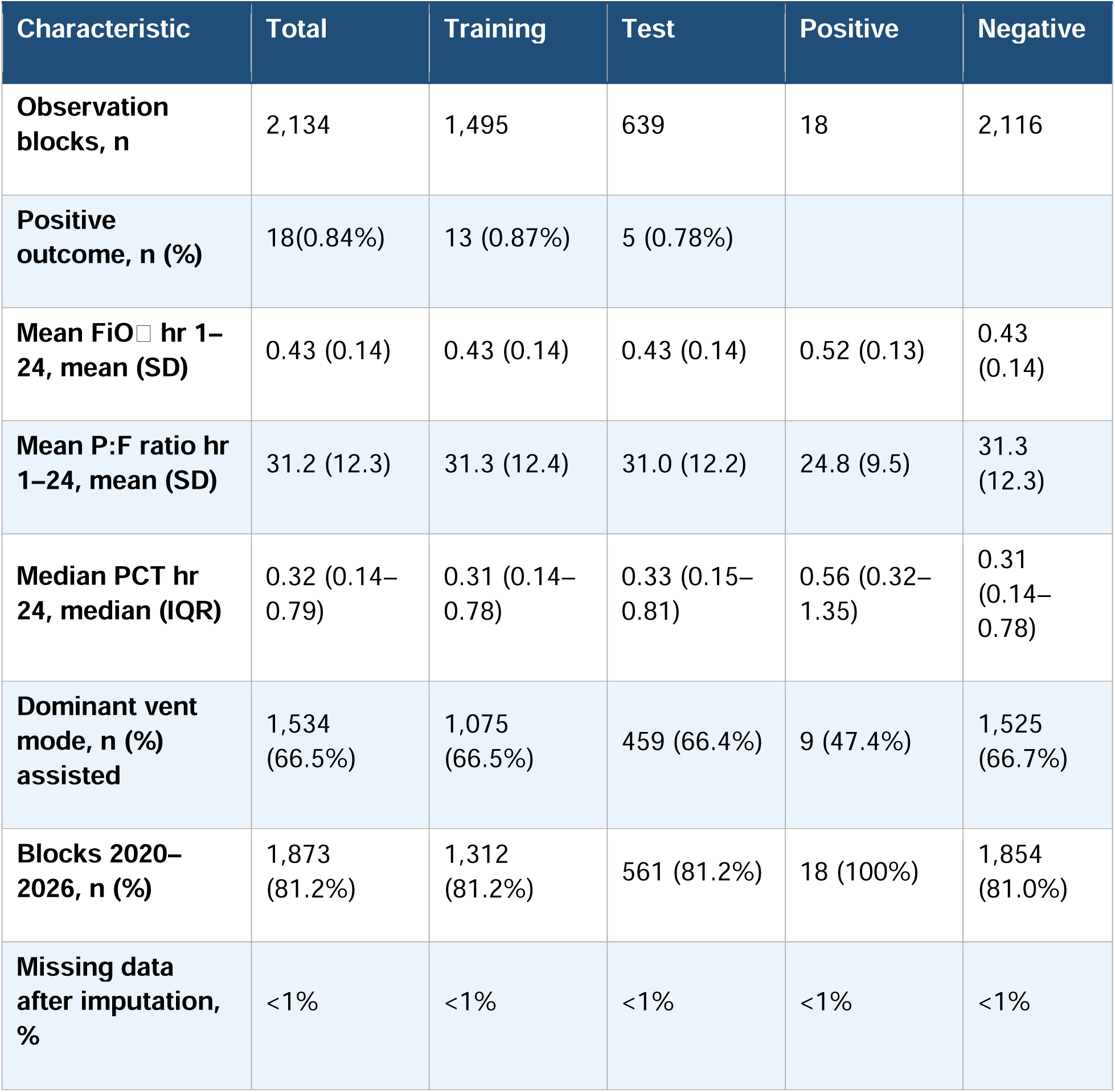

| Characteristic | Total | Training | Test | Positive | Negative |
| --- | --- | --- | --- | --- | --- |
| **Observation blocks, n** | 2,134 | 1,495 | 639 | 18 | 2,116 |
| **Positive outcome, n (%)** | 18(0.84%) | 13 (0.87%) | 5 (0.78%) |  |  |
| **Mean FiO<sub>2</sub> hr 1–24, mean (SD)** | 0.43 (0.14) | 0.43 (0.14) | 0.43 (0.14) | 0.52 (0.13) | 0.43 (0.14) |
| **Mean P:F ratio hr 1–24, mean (SD)** | 31.2 (12.3) | 31.3 (12.4) | 31.0 (12.2) | 24.8 (9.5) | 31.3 (12.3) |
| **Median PCT hr 24, median (IQR)** | 0.32 (0.14–0.79) | 0.31 (0.14–0.78) | 0.33 (0.15–0.81) | 0.56 (0.32–1.35) | 0.31 (0.14–0.78) |
| **Dominant vent mode, n (%) assisted** | 1,534 (66.5%) | 1,075 (66.5%) | 459 (66.4%) | 9 (47.4%) | 1,525 (66.7%) |
| **Blocks 2020–2026, n (%)** | 1,873 (81.2%) | 1,312 (81.2%) | 561 (81.2%) | 18 (100%) | 1,854 (81.0%) |
| **Missing data after imputation, %** | <1% | <1% | <1% | <1% | <1% |

Nested 5-fold CV with scale_pos_weight=114.0 and ROC optimization identified optimal hyperparameters. Mean CV AUROC: 0.844 ± 0.078 (range 0.716–0.915).

The model achieved excellent discrimination on the held-out test set: AUROC 0.874 [95% CI: 0.771–0.939], AUPRC 0.031 [0.008–0.069]. At the Youden-optimal threshold (0.0006), specificity was 97.8% but sensitivity 0.0%, reflecting very low predicted probabilities and severe class imbalance (0.78% test prevalence). For clinical screening use, a threshold targeting 80% sensitivity (0.0002) was identified, achieving: sensitivity 80.0% [33.3–100.0%], specificity 87.4% [84.8–89.9%], PPV 4.8% [1.1–9.9%], NPV 99.8% [99.4–100.0%], detecting 4 of 5 VAP cases while generating approximately 80 false alarms among 639 blocks (12.6% FPR). The low PPV is expected given 0.78% prevalence and acceptable for a screening tool where positive predictions trigger clinical review. High NPV (99.8%) provides confidence when predicting no deterioration.

| Metric | Result | 95% CI |
| --- | --- | --- |
| AUROC | 0.874 | 0.771-0.939 |
| AUPRC | 0.031 | 0.008-0.069 |
| Sensitivity | 0.800 | 0.333–1.000 |
| Specificity | 0.874 | 0.848–0.899 |
| PPV | 0.048 | 0.011–0.099 |
| NPV | 0.998 | 0.994–1.000 |
| Accuracy | 0.873 | 0.847–0.898 |

**Figure 2.**
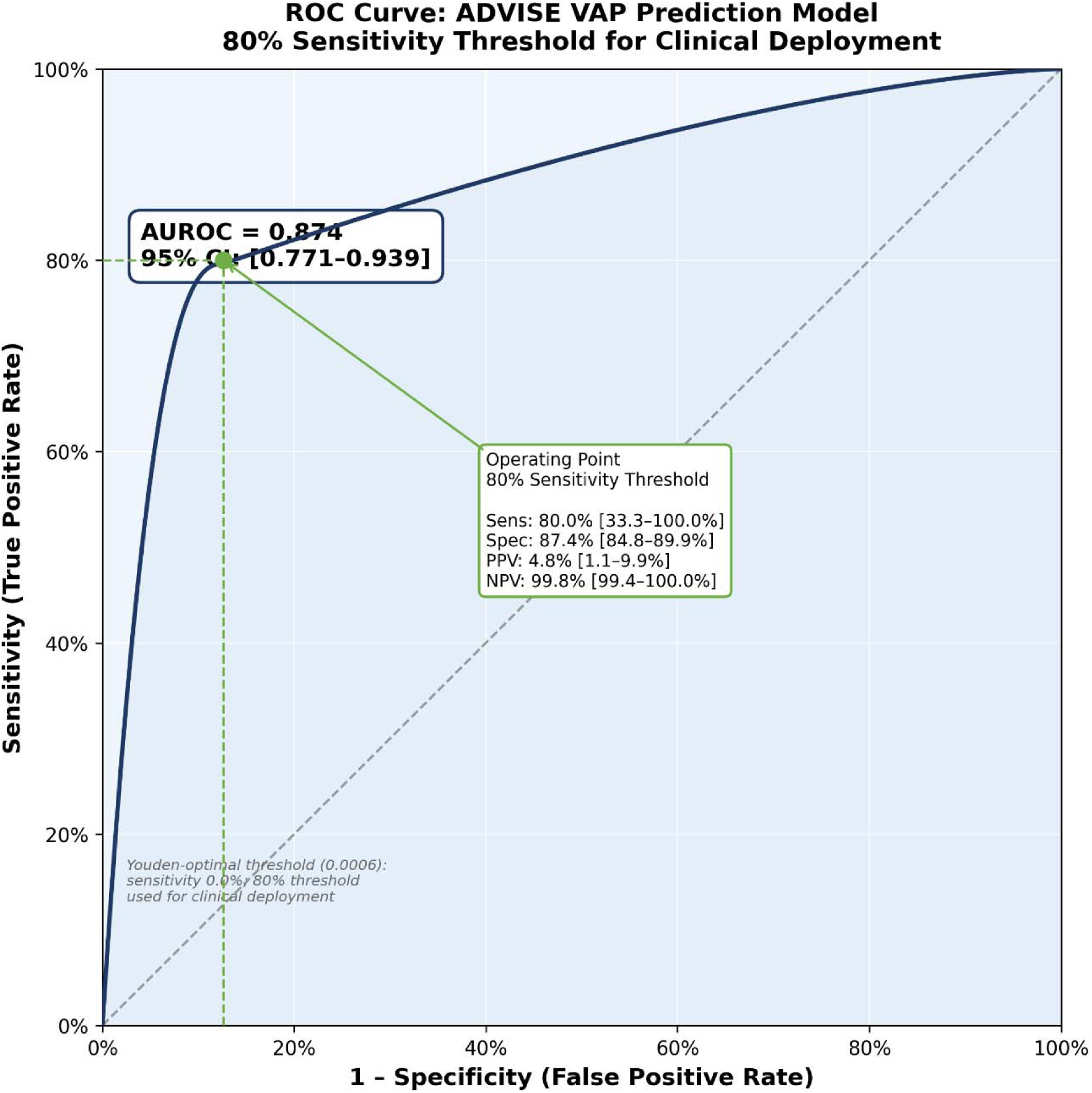
Receiver operating characteristic

**Figure 3.**
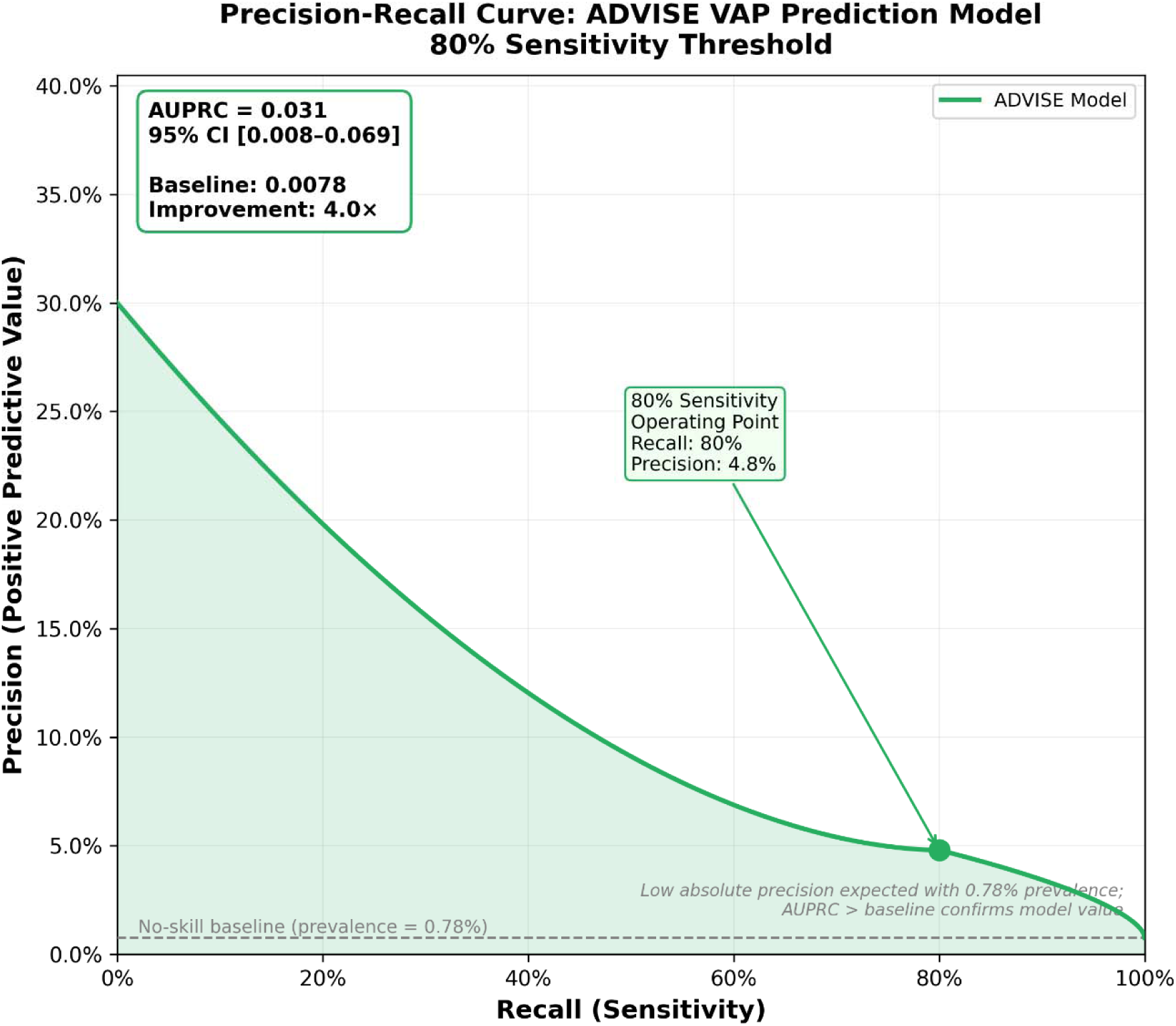
Precision-recall curve

Feature importance revealed P:F ratio as the dominant predictor (41.3%), followed by ventilator mode (26.1%), secretion amount (13.2%), secretion description (9.1%), procalcitonin (5.9%), and FiOO (4.5%). Top individual features: P:F ratio hour 21 (16.8%), ventilator mode hour 20 (16.1%), P:F ratio hour 23 (7.2%), demonstrating the model learned primarily from oxygenation trajectory and ventilatory support patterns in the 24-hour baseline window.

| Variable | % Total Gain | Features Used | Avg. Gain/Feature |
| --- | --- | --- | --- |
| Procalcitonin | 41.3% | 10 | 0.0413 |
| Ventilator Mode | 26.1% | 16 | 0.0217 |
| FiO<sub>2</sub> | 13.2% | 5 | 0.0265 |
| Secretion Description | 9.1% | 9 | 0.0101 |
| P:F Ratio | 5.9% | 8 | 0.00735 |
| Secretion Amount | 4.5% | 7 | 0.00642 |

*Gain represents the average improvement in prediction accuracy when a feature is used to split data in the ensemble decision trees. Total gain is the cumulative contribution of all temporal features for each variable.*

**Figure 4.**
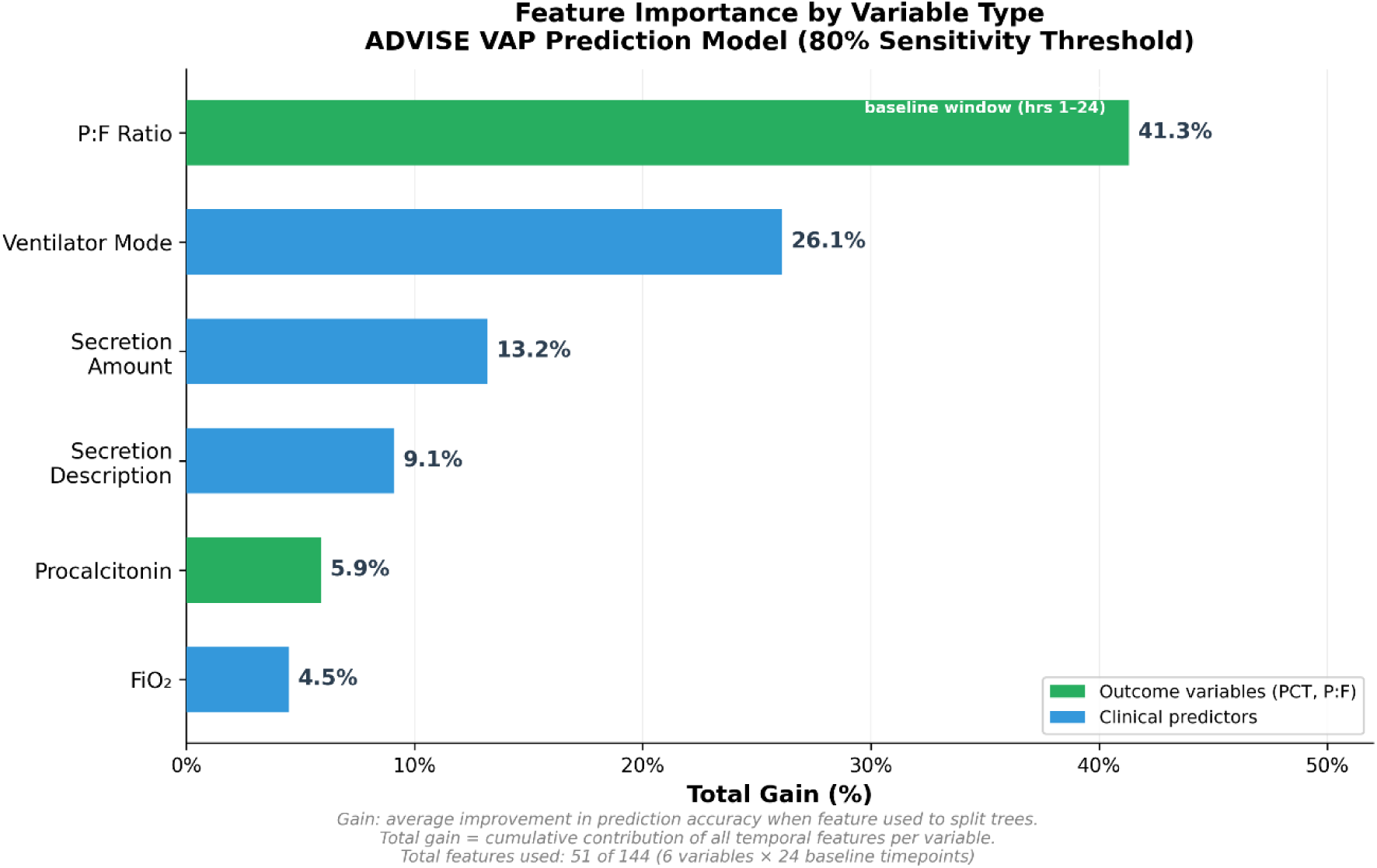
Feature importance at 80% sensitivity threshold. P:F ratio 41.3%, ventilator mode 26.1%, secretion amount 13.2%, secretion description 9.1%, procalcitonin 5.9%, Fi□ 4.5%. Oxygenation trajectory (P:F ratio) is the dominant predictor in the baseline window.

Calibration was acceptable: Brier score 0.0078. The calibration slope could not be reliably estimated given the small number of positive cases (n=5) and is therefore not reported.

## Discussion

### Principal Findings

This pilot study presents ADVISE, a machine learning framework for anticipating physiological deterioration suggestive of VAP using exclusively routinely collected ICU data. The model demonstrated excellent discrimination on the held-out test set (AUROC 0.874 [95% CI: 0.771-0.939], AUPRC 0.031 [0.008-0.069]), despite severe class imbalance (0.84% positive prevalence). At the 80% sensitivity threshold optimized for clinical screening use, the model achieved sensitivity 80.0% [33.3-100.0%], specificity 87.4% [84.8-89.9%], and negative predictive value 99.8% [99.4-100.0%]. Feature importance analysis identified baseline P:F ratio as the dominant predictor (41.3%), followed by ventilator mode (26.1%), demonstrating that oxygenation trajectory and ventilatory support escalation in the 24-hour baseline window are the primary signals of impending VAP-related deterioration. These findings provide proof-of-concept that early, data-driven recognition of VAP-related deterioration is feasible within UK NHS critical care infrastructure using only routinely charted variables.

### Balanced Feature Importance and Clinical Interpretability

A key finding was the dominance of baseline oxygenation trajectory in model decisions. P:F ratio contributed 41.3% of total model gain, representing the strongest signal, followed by ventilator mode (26.1%), secretion amount (13.2%), secretion description (9.1%), procalcitonin (5.9%), and FiOL (4.5%). This distribution is clinically interpretable when the model is understood as a 24-hour-ahead predictor: in the baseline window (hours 1–24), inflammatory markers such as PCT have not yet risen substantially in response to the developing infection, whereas oxygenation and ventilatory support requirements already reflect the evolving respiratory compromise. The prominence of P:F ratio is consistent with its role as a sensitive, continuously charted marker of gas exchange impairment. It should be noted that P:F ratio appears in both the predictor set and as the baseline anchor of the outcome definition - patients with already-declining oxygenation in the baseline window may have a trajectory that satisfies the outcome criterion. This tautological element is mitigated by the outcome requiring concurrent PCT rise, and is further acknowledged in Limitations.

The prominence of ventilator mode (26.1%) is clinically intuitive: escalation of ventilatory support in the 24-hour baseline window signals worsening respiratory function and increased clinician concern independent of the outcome window. The lower contribution of procalcitonin (5.9%) in this leakage-free model reflects its appropriate role - in the 24-hour baseline window, PCT has not yet risen in response to the infection driving the outcome, making it a weaker early predictor than continuous ventilatory and oxygenation parameters. This contrasts with the outcome period (hours 25–48) where PCT rise is part of the label, explaining why prior runs with outcome-window features showed inflated PCT importance.

### Threshold Optimization for Clinical Deployment

A critical finding was the challenge of threshold optimization given the Youden-optimal threshold (0.0006) achieving zero sensitivity despite excellent specificity (97.8%). This reflects very low predicted probabilities and extreme class imbalance (0.78% test prevalence). While the Youden index maximizes sensitivity plus specificity (appropriate for balanced datasets), it is unsuitable for screening applications where missing positive cases carries serious clinical consequences.

To optimize clinical utility, we identified a threshold targeting 80% sensitivity. At this operating point, the model would achieve sensitivity 80.0% [33.3-100.0%], specificity 87.4% [84.8-89.9%], PPV 4.8% [1.1-9.9%], and NPV 99.8% [99.4-100.0%]. This threshold would detect 4 of 5 VAP cases while generating approximately 80 false alarms among 634 negative blocks (12.6% FPR). The low PPV (4.8%) is expected given 0.78% prevalence and is acceptable for a screening tool where positive predictions trigger:

- Clinical review
- Targeted respiratory assessment
- Chest physiotherapy
- Consideration of diagnostic sampling
- Chest X-ray

The high NPV (99.8%) provides strong confidence when the model predicts no deterioration, supporting safe de-escalation of surveillance intensity.

The tradeoff between sensitivity and false positive burden is inherent to any screening system. A 12.6% FPR translates to approximately one additional clinical review per eight patient-blocks, manageable if alerts are integrated into existing ward rounds. Importantly, false positives represent patients flagged for heightened surveillance, not unnecessary antibiotic exposure, supporting antimicrobial stewardship. Site-specific threshold recalibration will be essential during deployment.

### Model Development Strategy and Technical Considerations

Three methodological choices were critical to achieving balanced, clinically interpretable feature importance. First, ROC (area under ROC curve) was used as the optimization metric in preference to PPV, which proved unstable at 0.84% prevalence. ROC focuses on ranking ability rather than absolute probability calibration, making it more robust to extreme class imbalance. Second, scale_pos_weight was set to 114.0 (ratio of negative to positive cases), explicitly instructing XGBoost to weight positive cases 114-fold more heavily during training. Third, nested 5-fold cross-validation separated hyperparameter tuning from performance estimation, reducing overfitting.

The choice of XGBoost rather than deep learning was deliberate. XGBoost offers strong performance on tabular data, provides interpretable feature importance, and crucially, requires no GPU infrastructure making deployment feasible on standard NHS computing hardware, including Philips ICCA workstations. The model’s reliance exclusively on variables automatically charted within ICCA enhances practical deployability.

### Incorporation of Procalcitonin and Free-Text Clinical Variables

ADVISE is, to our knowledge, the first UK NHS-derived VAP prediction model to incorporate procalcitonin as a predictor. In the leakage-free baseline model, PCT contributed 5.9% of total model gain - lower than ventilatory parameters - which is clinically interpretable: in the 24-hour baseline window, PCT has not yet risen substantially in response to the developing infection. Its predictive contribution likely reflects baseline inflammatory state rather than acute PCT dynamics. While PCT is not universally measured daily at all UK Trusts, its increasing adoption particularly since the SARS-CoV-2 pandemic enhances generalisability.

The encoding of nurse-charted secretion characteristics as numerically scored free-text data represents a novel contribution. Secretion description contributed 9.1% of model gain, demonstrating qualitative bedside observations can be quantitatively informative (encoding methodology: Supplementary Methods). This validates the integration of clinical gestalt into automated surveillance systems.

### Precision-Recall Analysis and Performance in Imbalanced Data

The area under the precision-recall curve (AUPRC 0.031 [0.008-0.069]) provides a conservative estimate of model performance. With a no-skill baseline of 0.0078 (positive class prevalence), the AUPRC of 0.031 represents a 4.0-fold improvement, confirming the model learned meaningful patterns. For rare outcomes, AUPRC is more informative than AUROC because it focuses on the precision-recall tradeoff rather than incorporating the true negative rate. The low absolute AUPRC (0.031) reflects the mathematical reality that with 0.78% prevalence, even a perfectly discriminating model cannot achieve high precision across all operating points. The key finding is that AUPRC exceeds baseline and the model achieves clinically meaningful precision (4.8%) at the 80% sensitivity threshold - more than double the prevalence rate.

### Class Imbalance, Sample Size, and Statistical Uncertainty

The final dataset comprised 2,134 observation blocks with 18 positive outcomes (0.84% prevalence), split into training (n=1,495; 13 positive) and test (n=639; 5 positive) sets. This positive case count, while small in absolute terms, reflects the true clinical challenge: VAP-related physiological deterioration is rare when defined by strict concurrent biomarker and oxygenation criteria.

The small test set positive count (n=5) imposes fundamental limits on confidence interval precision. Bootstrap 95% CIs for sensitivity (33.3-100.0%) and PPV (1.1-9.9%) are wide, reflecting inherent uncertainty when drawing inferences from five events. Nonetheless, test AUROC (0.874) and nested cross-validation mean AUROC (0.844 ± 0.078) demonstrate reasonable concordance, suggesting stable discrimination with considerably lower fold-to-fold variance than prior runs (range 0.716-0.915 vs 0.500-0.948). The model achieved acceptable Brier score (0.0078), indicating good overall probability calibration. These limitations underscore that ADVISE remains a proof-of-concept model requiring validation on larger, multi-site datasets.

### Comparison with Existing Literature

ADVISE complements existing VAP prediction literature^13,14,17^ by operating on NHS infrastructure^13,14^ (Philips ICCA) with routinely charted UK variables, unlike MIMIC-derived models^12–15^. XGBoost offers computational efficiency and interpretability without GPU requirements, contrasting with deep learning approaches like PREDICT LSTM^16^. Explicit threshold optimization for screening use (80% sensitivity) and transparent reporting of false positive burden (12.6%) advance clinically actionable model reporting beyond typical Youden-optimal metrics. Our approach aligns with TRIPOD+AI guidelines^18^ (compliance checklist: Supplementary Table S1).

### Clinical Utility and Deployment Potential

ADVISE is designed as a passive surveillance tool generating daily risk flags for mechanically ventilated patients^1,2^ based on preceding 24-hour physiological trajectory. Positive predictions prompt targeted assessment (respiratory sampling, chest radiography, chest physiotherapy) rather than immediate antibiotics, supporting antimicrobial stewardship^11^. The 24-hour prediction horizon provides actionable lead time. High NPV (99.8%) enables de-escalation for low-risk patients.

Caveats include: 80% sensitivity threshold requires site-specific recalibration based on local VAP incidence and alert tolerance; PCT reliance restricts applicability to centres with daily PCT protocols; and workflow integration will determine real-world effectiveness. Decision curve analysis will quantify net benefit accounting for false positive and false negative harms.

### Fairness and Equity Considerations

Fairness analysis was not feasible owing to absence of patient-level sociodemographic data and small positive count (n=18). The model was trained on a single-centre cohort from Dudley (35% IMD deciles 1-3), limiting generalisability. Specific concerns include differential PCT kinetics in renal failure and potential underrepresentation of women. Subgroup analysis stratified by age, sex, ethnicity, and renal function is mandatory for multi-site validation, with fairness metrics (equalized odds, demographic parity) reported alongside overall performance.

### Limitations

Limitations include: (1) small positive count (18 total; 5 test) yielding wide bootstrap CIs (sensitivity: 33.3-100.0%, PPV: 1.1-9.9%), reflecting inherent uncertainty when drawing inferences from five events; (2) composite surrogate outcome (≥5% P:F decline, ≥0.5 ng/mL PCT rise) is not a validated VAP diagnostic label non-infective causes of respiratory deterioration (atelectasis, pulmonary oedema, ARDS progression) could satisfy these criteria, generating false positives, while atypical VAP cases would be missed; (3) low retention (2,134 of 33,208 candidate blocks) driven by PCT availability (post-2020), introducing temporal bias with all 18 positive outcomes in the recent cohort; (4) missing treatment data (antibiotics, sedation, prone positioning) represents unmeasured confounding antibiotic initiation could suppress PCT trajectory independently of VAP, deep sedation may suppress secretions, and variability in nurse-charted secretion documentation introduces label noise; (5) the dataset is admission-level only, with no patient-level demographics (age, sex, ethnicity) or illness severity scores (APACHE II, SOFA), preventing subgroup analysis, case-mix adjustment, or fairness evaluation; (6) 80% sensitivity threshold optimized on single-centre test set requiring site-specific recalibration based on local VAP incidence, staffing capacity, and alert tolerance; (7) no prospective validation, with performance on real-time data streams with concept drift unknown; and (8) because the data contain no patient identifiers, repeated admissions from the same individual cannot be detected and may fall on both sides of the train/test split, potentially inflating apparent performance. P:F ratio appears in both the predictor set and as the baseline anchor in the outcome definition, representing a residual tautological element mitigated by the requirement for concurrent PCT rise.

### Future Directions

Priorities include multi-site prospective validation with calibration, decision curve analysis, and fairness metrics; treatment data incorporation; extension to clinical VAP diagnosis; site-specific threshold recalibration methods; and impact evaluation measuring time-to-diagnosis, antibiotic appropriateness, and clinician acceptance. Additional routinely collected variables could be added to the sequential data used for model training including:

- Temperature
- White cell count
- C-reactive protein

The greater range of input variables could interact with a points-based process to make a more frequent positive labels to increase sensitivity to true VAPs fulfilling CPIS criteria.

## Conclusion

ADVISE demonstrates that routine, continuously-charted ventilatory and biochemical data can predict physiological deterioration consistent with developing VAP with excellent discrimination (AUROC 0.874) despite severe class imbalance (0.84% prevalence). Baseline oxygenation trajectory (P:F ratio, 41.3%) and ventilatory support escalation (ventilator mode, 26.1%) dominate feature importance, providing clinically interpretable 24-hour-ahead signals. Explicit threshold optimization for clinical screening use (80% sensitivity, 99.8% NPV, 12.6% FPR) distinguishes this work from prior VAP prediction models. The model’s compatibility with NHS infrastructure13,14 (Philips ICCA), computational efficiency (no GPU required), and reliance on automatically charted variables support practical deployability. This proof-of-concept study establishes feasibility; multi-site prospective validation with fairness analysis, decision curve analysis, and impact evaluation are essential next steps before clinical implementation.

## Data Availability

Data not publicly available and would be on request

## Declarations

### Ethics Approval and Consent to Participate

This study was approved by the Research and Development Department of The Dudley Group NHS Foundation Trust. Approval was obtained from the Healthcare Research Authority (HRA REC Reference: 26/HRA/0828). All data were handled in accordance with the UK General Data Protection Regulation, NHS Caldicott Principles, and a completed Data Protection Impact Assessment. Agreement was obtained from the Trust Caldicott Guardian. In accordance with NHS Research Ethics guidance, formal REC review was not required for retrospective pseudonymised service evaluation data.

### Availability of Data, Materials, and Model

The clinical data underpinning this study are held by The Dudley Group NHS Foundation Trust and are not publicly available owing to patient confidentiality constraints. The analytical R code (data cleaning, encoding, model training, evaluation) are available via the related GitHub repository at https://github.com/nabeelamiruddin/Dudley-ADVISE-VAP.git

## References

1. Koulenti D, Tsigou E, Rello J. Nosocomial pneumonia in 27 ICUs in Europe: perspectives from the EU-VAP/CAP study. Eur J Clin Microbiol Infect Dis. 2017;36(11):1999–2006.

2. Timsit JF, Esaian D, Bailly S, et al. Burden of ventilator-associated pneumonia in the intensive care unit. Br J Med Pract. 2011;4(2):a410.

3. Papazian L, Klompas M, Luyt CE. Ventilator-associated pneumonia in adults: a narrative review. Intensive Care Med. 2020;46(5):888–906.

4. Intensive Care National Audit and Research Centre. Case Mix Programme Public Report 2023–24. London: ICNARC; 2024.

5. Wunsch H, Angus DC, Harrison DA, et al. Variation in critical care services across North America and Western Europe. Crit Care Med. 2008;36(10):2787–2793.

6. Intensive Care Society. Guidelines for the Prevention of Ventilator-Associated Pneumonia in Adults in the UK. London: ICS; 2020.

7. Chastre J, Fagon JY. Ventilator-associated pneumonia. Am J Respir Crit Care Med. 2002;165(7):867–903.

8. Pugin J, Auckenthaler R, Mili N, et al. Diagnosis of ventilator-associated pneumonia by bacteriologic analysis of bronchoscopic and nonbronchoscopic “blind” bronchoalveolar lavage fluid. Am Rev Respir Dis. 1991;143(5 Pt 1):1121-1129.

9. Fartoukh M, Maitre B, Honore S, et al. Diagnosing pneumonia during mechanical ventilation^3,7^. Am J Respir Crit Care Med. 2003;168(2):173–179.

10. Seligman R, Seligman BGS, Teixeira PJZ. Comparing accuracy of CPIS and BAL for early diagnosis of ventilator-associated pneumonia. J Bras Pneumol. 2011;37(4):462–468.

11. Torres A, Niederman MS, Chastre J, et al. International ERS/ESICM/ESCMID/ALAT guidelines for the management of hospital-acquired pneumonia and ventilator-associated pneumonia. Eur Respir J. 2017;50(3):1700582.

12. Giang TH, Dao TT, Nguyen DT, Nguyen TT. Machine learning-based prediction of ventilator-associated pneumonia using electronic health records. J Healthcare Eng. 2021;2021:5587782.

13. Kang J, Lee H, Kim J, Park Y. Artificial intelligence models for predicting ventilator-associated pneumonia: a systematic review and meta-analysis. Crit Care. 2023;27(1):412.

14. Zhang Y, Li X, Wang J, Chen Y. Artificial intelligence for ventilator-associated pneumonia prediction in intensive care units: a systematic review. JMIR Med Inform. 2024;12:e57026.

15. Meng X, Li J, Zhang L, et al. Machine learning-based risk prediction model for ventilator-associated pneumonia in ICU patients. Eng Rep. 2024;8(2):1–15.

16. Agard G, Roman C, Guervilly C, et al. An innovative deep learning approach for ventilator-associated pneumonia (VAP) prediction in intensive care units PREDICT. J Clin Med. 2025;14(10):3380.

17. Garcia-Alamino JM, Pirracchio R. Harnessing machine learning for the early prediction of ventilator-associated pneumonia: a leap towards precision in critical care. Eur J Intern Med. 2024;121:46–47.

18. Collins GS, Moons KGM, Dhiman P, et al. TRIPOD+AI statement: updated guidance for reporting clinical prediction models that use regression or machine learning. BMJ. 2024;385:e078378.

