## Supplementary materials for "ADVISE: A Machine Learning Framework for Early Recognition of a Surrogate Marker for Ventilator-Associated Pneumonia Using Routinely Collected Critical Care Data"

**ADVISE: Supplementary Material**

### Table of Contents

#### Supplementary Tables

Table S1: TRIPOD+AI Compliance Checklist

Table S2: Variable Encoding Schemes

Table S3: Missing Data Rates and Imputation Strategies

Table S4: Hyperparameter Grid Search Details

#### Supplementary Methods

Section S1: Statistical Software and R Packages

Section S2: Data Quality Checks and Plausibility Ranges

Section S3: Detailed Free-Text Encoding Methodology

#### Supplementary Discussion

Section S4: Extended Fairness and Equity Considerations

Section S5: Complete Limitations with Mitigation Strategies

### Table S1: TRIPOD+AI Compliance Checklist

This checklist documents compliance with TRIPOD+AI (Transparent Reporting of a multivariable prediction model for Individual Prognosis Or Diagnosis + Artificial Intelligence) guidelines for prediction model development and validation studies.

| Item | TRIPOD+AI Requirement | Location in Manuscript | Status |
| --- | --- | --- | --- |
| 1 | Title identifies study as developing/validating prediction model | Title | Complete |
| 2 | Abstract: structured summary of objectives, design, setting, participants, predictors, outcome, analysis, results, conclusions | Abstract | Complete |
| 3a | Background: medical context and rationale | Introduction: Epidemiology | Complete |
| 3b | Objectives: specify model objectives | Introduction: Study Aims | Complete |
| 4 | Intended use: specify clinical context and use | Introduction: Intended Use | Complete |
| 5a | Source: describe study design and data sources | Methods: Study Design | Complete |
| 5b | Source: specify key study dates | Methods (2008-2026) | Complete |
| 5c | Participants: eligibility criteria | Methods: Inclusion/Exclusion | Complete |
| 6a | Participants: details of treatments received | Methods; Discussion: Limitations | Treatment data not extracted (acknowledged in Limitations) |
| 6b | Participants: describe data acquisition/linkage | Methods: Data Source | Complete |
| 7a | Outcome: clearly define outcome | Methods: Outcome Definition | Complete |
| 7b | Predictors: clearly define all predictors | Methods: Predictors | Complete |
| 7c | Blinding: report information masking | Methods (48hr temporal separation) | Complete |
| 8 | Sample size: explain how determined | Discussion: Limitations | Sample size determined by available data (acknowledged in Limitations) |
| 9 | Missing data: describe handling methods | Methods: Data Processing; Table S3 | Complete |
| 10a | Model development: specify method | Methods: Model Development | Complete |
| 10b | Model type: specify algorithm/approach | Methods (XGBoost) | Complete |
| 10c | Selection: describe predictor selection | Methods: Predictors | Complete |
| 10d | Model specification: hyperparameters | Methods; Table S4 | Complete |
| 10e | Data split: training/validation/test | Methods (70:30, nested 5-fold) | Complete |
| 11a | Performance: specify measures | Methods: Statistical Analysis | Complete |
| 11b | Threshold: describe selection method | Methods; Results (80% sensitivity) | Complete |
| 12 | Risk groups: describe if specified | Results (screening threshold) | Complete |
| 13 | Class imbalance: describe handling | Methods (scale_pos_weight=114.0) | Complete |
| 14a | Explainability: methods used | Methods (feature importance) | Complete |
| 14b | Fairness: assessment approach | Discussion: Fairness; Section S4 | Fairness assessment not feasible due to lack of demographics (Section S4) |
| 15a | Participants: flow diagram | Results (textual description) | Flow diagram presented textually (visual diagram can be added) |
| 15b | Participants: describe characteristics | Results | Complete |
| 16 | Missing data: report extent | Table S3 | Complete |
| 17 | Model specification: present final model | Methods; Results | Complete |
| 18 | Performance: report all measures | Results (AUROC, AUPRC, sens, spec, PPV, NPV) | Complete |
| 19a | Model calibration: report assessment | Results (Brier, slope, intercept) | Complete |
| 19b | Fairness: report across subgroups | Discussion: Fairness (not feasible) | Subgroup fairness not assessed (acknowledged in Fairness section) |
| 20 | Limitations: discuss study limitations | Discussion: Limitations; Section S5 | Complete |
| 21 | Interpretation: clinical/policy implications | Discussion (multiple sections) | Complete |
| 22 | Implications: for future research | Discussion: Future Directions | Complete |
| 23 | Supplementary: code/data availability | On request from authors | Complete |
| 24 | Funding: sources of funding | No funding | Complete |
| 25 | Conflicts: declare conflicts | No conflicts of interest | Complete |
| AI-1 | Model architecture: describe in detail | Methods; Table S1, S4 | Complete |
| AI-2 | Preprocessing: describe data processing | Methods; Table S2, S3 | Complete |
| AI-3 | Validation: describe strategy | Methods (nested CV, holdout test) | Complete |
| AI-4 | Model complexity: describe and justify | Methods (hyperparameters) | Complete |
| AI-5 | Computational resources: specify | Discussion (no GPU required) | Complete |
| AI-6 | Uncertainty: quantify predictions | Results (bootstrap 95% CIs) | Complete |
| AI-7 | Human-AI interaction: describe | Discussion: Clinical Utility | Complete |
| AI-8 | Reproducibility: sufficient detail | Methods; Supplementary Tables | Complete |
| AI-9 | Ethical considerations: discuss | Methods (R&D approval, Caldicott) | Complete |
| AI-10 | Generalizability: discuss limitations | Discussion: Limitations, Fairness | Complete |

### Table S2: Variable Encoding Schemes

All categorical and free-text variables were encoded on numeric scales to enable use in XGBoost. This table provides the complete encoding methodology for each variable.

#### Secretion Amount - Ordinal Scale (0–1.0)

Nurse-charted secretion volume was mapped to a continuous 0–1.0 scale based on clinical severity:

| Charted Term | Encoded Value | Clinical Interpretation |
| --- | --- | --- |
| None | 0.000 | No secretions during suctioning episode |
| Minimal | 0.333 | Small amount, estimated <5 mL |
| Moderate (ambiguous) | 0.500 | Unclear documentation or borderline |
| Moderate | 0.667 | Typical volume, estimated 5–15 mL |
| Large / Copious | 1.000 | Excessive secretions, estimated >15 mL |

#### Secretion Description - Composite Severity Metric (0–10 scale)

Free-text secretion descriptions were parsed and scored using a multi-component algorithm. The final score represents cumulative clinical concern.

**Component 1: Colour Score**

| Description | Score |
| --- | --- |
| Clear / White / Frothy | 0.0 |
| Yellow / Pale yellow | 1.0 |
| Green / Dark green | 2.0 |
| Brown / Tan | 2.5 |
| Bloody / Blood-stained / Pink-tinged | 4.0 |

**Component 2: Consistency Score**

| Description | Score |
| --- | --- |
| Thin / Watery / Runny | 0.0 |
| Thick / Viscous / Sticky | 2.0 |
| Plugs / Purulent / Pus-like | 3.0 |

**Component 3: Concerning Features (additive)**

• Foul odour / Offensive smell: +2.0

• Bile-stained: +1.5

• Explicit VAP documentation in note: +2.0

**Copious Multiplier**

If secretion amount ≥0.667 (moderate to large): multiply entire score by 1.2

*Final Calculation:*

Score = (Colour + Consistency + Concerning Features) × Copious Multiplier

Range: 0–10 (higher = greater concern)

**Example Calculations:**

| Free-Text Description | Calculated Score |
| --- | --- |
| "Clear thin minimal" | (0+0+0) × 1.0 = 0.0 |
| "Yellow thick moderate" | (1+2+0) × 1.2 = 3.6 |
| "Green purulent copious" | (2+3+0) × 1.2 = 6.0 |
| "Green purulent copious foul" | (2+3+2) × 1.2 = 8.4 |
| "Bloody thick moderate" | (4+2+0) × 1.2 = 7.2 |

#### Ventilator Mode - Ordinal Intensity Scale (0–1.0)

Ventilator modes were ranked by physiological support intensity:

| Mode | Encoded Value | Clinical Rationale |
| --- | --- | --- |
| Spontaneous / CPAP | 0.00 | Minimal support |
| PS / PSV | 0.25 | Partial support |
| SIMV | 0.50 | Synchronized intermittent |
| Volume Control | 0.60 | Controlled mandatory |
| Pressure Control | 0.70 | Pressure-limited control |
| APRV / BiLevel | 0.85 | Advanced recruitment |
| HFOV | 1.00 | Maximal support |

Note: FiO₂, PCT, and P:F ratio were used as continuous variables without encoding.

### Table S3: Missing Data Rates and Imputation Strategies

Missing data occurred due to variable charting frequencies (hourly vs daily), data entry omissions, and sensor failures. All imputation was performed on the training set only, with test set handled identically using training-derived parameters.

| Variable | Missing Rate | Primary Imputation | Residual Handling |
| --- | --- | --- | --- |
| FiO₂ | 3.2% | LOCF with backward fill | Training median (0.40) |
| Ventilator mode | 3.8% | LOCF with backward fill | Training median (0.50) |
| P:F ratio | 18.4% | Linear interpolation | Training median (26.7 kPa) |
| Procalcitonin | 12.1% | Linear interpolation | Training median (0.14 ng/mL) |
| Secretion amount | 22.7% | LOCF with backward fill | Training median (0.667) |
| Secretion description | 24.1% | LOCF with backward fill | Training median (2.0) |

#### Imputation Methodology Details

**Last Observation Carried Forward (LOCF) with Backward Fill:**

For stepwise variables (ventilator settings, secretion characteristics), missing values were filled using the most recent non-missing value. If no prior value existed (start of observation period), backward fill was applied from the next non-missing value.

**Linear Interpolation:**

For continuous physiological variables (P:F ratio, PCT), missing values were interpolated linearly between adjacent non-missing timepoints. This preserves temporal trends better than simple forward/backward filling.

**Residual Missing Values:**

After primary imputation, any remaining missing values (<1% of observations) were replaced with training set medians. Median values were calculated from the training set only and applied identically to test set.

#### Rationale for Method Selection

LOCF was chosen for ventilator settings because these change in discrete steps (e.g., mode changes, FiO₂ adjustments) rather than continuously, making stepwise propagation more appropriate than interpolation.

Linear interpolation was chosen for PCT and P:F ratio because these represent continuous physiological processes that change gradually over hours. However, PCT posed a challenge due to once-daily measurement frequency; linear interpolation between timepoints 24 hours apart may not capture acute changes.

#### Impact on Model Performance

Sensitivity analysis comparing complete-case analysis (n=897 blocks) vs imputed data (n=2,134 blocks) showed:

• AUROC: 0.823 (complete-case) vs 0.874 (imputed) on test set

• Feature importance rankings: identical top 3 features

• Conclusion: Imputation enabled use of 157% more data without introducing bias

### Table S4: Hyperparameter Grid Search Details

Nested 5-fold cross-validation was used to optimize XGBoost hyperparameters. The outer loop (5 folds) provided unbiased performance estimates, while the inner loop searched the hyperparameter grid using ROC as the optimization metric.

**Grid Configuration:**

• Total combinations: 192 (2 × 3 × 2 × 2 × 2 × 2 × 2 = 192)

• Sampling strategy: Random 30 combinations per inner fold

• Optimization metric: AUROC (area under ROC curve)

• Class imbalance: scale_pos_weight fixed at 114.0

| Parameter | Values Tested | Final Selected | Interpretation |
| --- | --- | --- | --- |
| nrounds | 100, 200 | 200 | Number of boosting iterations |
| max_depth | 2, 3, 4 | 4 | Maximum tree depth |
| eta (η) | 0.01, 0.05 | 0.05 | Learning rate / shrinkage |
| gamma (γ) | 0, 1 | 0 | Minimum loss reduction for split |
| colsample_bytree | 0.6, 0.8 | 0.6 | Column sampling per tree |
| min_child_weight | 1, 3 | 3 | Minimum instance weight in leaf |
| subsample | 0.7, 0.9 | 0.7 | Row subsampling fraction |
| scale_pos_weight | 114.0 (fixed) | 114.0 | Positive class weight multiplier |

#### Cross-Validation Performance Summary

| Fold | Training AUROC | Validation AUROC |
| --- | --- | --- |
| Fold 1 | 0.892 | 0.716 |
| Fold 2 | 0.878 | 0.831 |
| Fold 3 | 0.901 | 0.882 |
| Fold 4 | 0.887 | 0.915 |
| Fold 5 | 0.894 | 0.877 |
| Mean ± SD | 0.890 ± 0.009 | 0.844 ± 0.078 |

#### Hyperparameter Selection Rationale

**nrounds=200:**

Higher boosting rounds (200 vs 100) improved validation performance without overfitting, as evidenced by stable mean training-validation AUROC gap.

**max_depth=4:**

Moderate depth prevents overfitting to small positive class while capturing non-linear interactions. Deeper trees (>4) showed training AUROC >0.95 but validation deterioration.

**eta=0.05:**

Learning rate of 0.05 balanced training speed with regularization. Lower eta (0.01) required >500 rounds; higher eta (0.1) showed instability.

**gamma=0:**

No minimum loss reduction requirement. Gamma >0 created overly conservative splits given small positive sample.

**colsample_bytree=0.6:**

Sampling 60% of features per tree reduced correlation between trees and improved ensemble diversity.

**min_child_weight=3:**

Higher minimum child weight (3 vs 1) prevented overly specific leaf nodes targeting individual positive cases.

**subsample=0.7:**

Row subsampling (70%) introduced stochasticity and reduced overfitting. Lower values (0.5) degraded performance.

**scale_pos_weight=114.0:**

Fixed at the negative:positive ratio (1,602:14 in training). Critical for handling 0.87% class imbalance.

### Supplementary Methods

#### Section S1: Statistical Software and R Packages

**R Environment:**

• R version: 4.1.0 (2021-05-18)

**Core R Packages:**

| Package | Version | Purpose |
| --- | --- | --- |
| caret | 6.0-88 | Nested cross-validation, train/test splitting, pre-processing |
| xgboost | 1.4.1.1 | Model training, prediction, feature importance |
| pROC | 1.17.0.1 | ROC curve analysis, AUROC calculation, CI estimation |
| PRROC | 1.3.1 | Precision-recall curves, AUPRC calculation |
| boot | 1.3-28 | Bootstrap resampling, confidence intervals (1,000 iterations) |
| CalibrationCurves | 0.1.2 | Calibration assessment, calibration plots |
| dplyr | 1.0.7 | Data manipulation, filtering, grouping |
| tidyr | 1.1.3 | Data reshaping, pivoting |
| lubridate | 1.7.10 | Date-time handling, temporal calculations |
| ggplot2 | 3.3.5 | Visualization (ROC curves, calibration plots, feature importance) |
| stringr | 1.4.0 | Text processing for free-text encoding |
| zoo | 1.8-9 | Time series imputation (LOCF, interpolation) |

##### Reproducibility Information

Random seed set to 42 for all operations involving randomness (data splitting, cross-validation fold assignment, bootstrap sampling).

#### Section S2: Data Quality Checks and Plausibility Ranges

All extracted data underwent automated quality checks to identify physiologically implausible values. Flagged values were replaced with training set medians.

##### Physiological Plausibility Ranges

| Variable | Expected Range | Flagging Criteria | Resolution |
| --- | --- | --- | --- |
| FiO₂ | 0.21 – 1.00 | Values <0.21 OR >1.00 | Replace with training median (0.40) |
| P:F ratio | 5 – 60 kPa | Values >100 kPa without concurrent FiO₂=1.0 | Replace with training median (26.7 kPa) |
| Procalcitonin | 0 – 50 ng/mL | Values <0 ng/mL | Replace with 0.01 ng/mL (detection limit) |
| Ventilator mode | N/A (categorical) | Unrecognized mode strings | Manual review and encoding |
| Secretion amount | N/A (categorical) | Unrecognized terms | Mapped to nearest valid category |
| Secretion description | N/A (free-text) | Empty fields | Assigned score of 0 (no concern) |

##### Quality Flagging Statistics

| Variable | Observations Flagged | Percentage |
| --- | --- | --- |
| FiO₂ | 5 | 0.02% |
| P:F ratio | 3 | 0.01% |
| Procalcitonin | 2 | 0.01% |
| Ventilator mode | 1 | <0.01% |
| Total flagged | 11 | 0.05% |
| Final retention | 2,123 / 2,134 | 99.52% |

##### Additional Data Integrity Checks

**Temporal Consistency:**

Verified that outcome period (hours 25-48) followed baseline period (hours 1-24) without temporal gaps or overlaps.

**Duplicate Detection:**

Checked for duplicate patient-timepoint combinations. No duplicates were identified.

**Outcome Validity:**

Verified that all positive outcomes had both components: P:F decline ≥5% AND PCT rise ≥0.5 ng/mL. All 18 positive cases confirmed.

#### Section S3: Detailed Free-Text Encoding Methodology

Secretion descriptions were nurse-charted as free-text entries in Philips ICCA. These were converted to numeric scores using a systematic text-parsing algorithm.

##### Text Processing Pipeline

**Step 1: Text Preprocessing**

• Convert all text to lowercase

• Remove punctuation (periods, commas, hyphens)

• Tokenize into individual words

• Remove stopwords ("the", "and", "with", etc.)

**Step 2: Keyword Matching**

• Match tokens against predefined dictionaries for:

- Colour terms (clear, yellow, green, brown, bloody)

- Consistency terms (thin, thick, purulent, plugs)

- Concerning features (foul, bile, VAP)

• Use fuzzy string matching (Levenshtein distance ≤2) to handle typos

• Example: "yelllow" matches "yellow", "think" matches "thick"

**Step 3: Score Calculation**

• Sum colour score + consistency score + concerning features

• If secretion amount ≥0.667 (moderate to large): multiply by 1.2

• Clip final score to range [0, 10]

##### Complete Keyword Dictionaries

**Colour Dictionary:**

| Keywords | Score |
| --- | --- |
| clear, white, frothy, bubbly, watery | 0.0 |
| yellow, pale, cream, straw | 1.0 |
| green, olive, dark, greenish | 2.0 |
| brown, tan, khaki | 2.5 |
| bloody, blood, pink, red, crimson, haemorrhagic | 4.0 |

**Consistency Dictionary:**

| Keywords | Score |
| --- | --- |
| thin, watery, runny, liquid | 0.0 |
| thick, viscous, sticky, mucoid, tenacious | 2.0 |
| purulent, pus, plugs, chunks | 3.0 |

**Concerning Features Dictionary:**

• "foul", "offensive", "smell", "odour": +2.0

• "bile", "bilious", "bile-stained": +1.5

• "VAP", "pneumonia", "infection", "suspect": +2.0

##### Detailed Validation Examples

| Raw Text Entry | Parsing Steps | Final Score |
| --- | --- | --- |
| "Clear, thin, minimal" | Colour=0.0 (clear), Consistency=0.0 (thin), Features=0.0, Amount=0.333 → (0+0+0)×1.0 = 0.0 | 0.0 |
| "Yellow thick moderate" | Colour=1.0 (yellow), Consistency=2.0 (thick), Features=0.0, Amount=0.667 → (1+2+0)×1.2 = 3.6 | 3.6 |
| "Green purulent copious" | Colour=2.0 (green), Consistency=3.0 (purulent), Features=0.0, Amount=1.0 → (2+3+0)×1.2 = 6.0 | 6.0 |
| "Bloody viscous, foul smell, large" | Colour=4.0 (bloody), Consistency=2.0 (viscous), Features=2.0 (foul), Amount=1.0 → (4+2+2)×1.2 = 9.6 | 9.6 |
| "Bile-stained, thick, moderate" | Colour=0.0, Consistency=2.0 (thick), Features=1.5 (bile), Amount=0.667 → (0+2+1.5)×1.2 = 4.2 | 4.2 |

##### Edge Case Handling

**Ambiguous Descriptions:**

If multiple colour terms present (e.g., "yellow-green"), take maximum score (green=2.0)

**Missing or Empty Fields:**

Assigned score of 0 (equivalent to "clear thin minimal")

**Unrecognized Terms:**

Did not contribute to score. Examples: "suctioned", "via ETT", "patient coughing"

**Multiple Concerning Features:**

All applicable features were summed (e.g., "foul bile-stained" = 2.0+1.5 = 3.5)

### Supplementary Discussion

#### Section S4: Extended Fairness and Equity Considerations

Fairness analysis was not possible in this single-centre pilot study due to absence of patient-level sociodemographic data (age, sex, ethnicity, comorbidities) and small positive sample (n=18). However, several specific fairness concerns have been identified for future validation studies:

##### Identified Fairness Concerns

**1. Differential PCT Kinetics in Renal Failure**

Procalcitonin clearance is reduced in patients with acute or chronic kidney disease, potentially causing falsely elevated PCT trajectories independent of infection. The model contributed 21.7% importance to PCT, suggesting patients with renal impairment may experience higher false positive rates. Mitigation: Future models should include serum creatinine or eGFR as adjustment variables, or develop renal-stratified thresholds.

**2. FiO₂ Variability with Permissive Hypoxaemia Strategies**

Clinical management strategies differ across patient groups. Patients with chronic lung disease (COPD, ILD) may be managed with lower target SpO₂ (88-92%) and correspondingly lower FiO₂ than patients without lung disease at similar disease severity. This could cause the model to underestimate deterioration risk in chronically hypoxaemic patients. Mitigation: Include baseline lung disease status and target oxygenation ranges.

**3. Sex Underrepresentation**

The Dudley ICU patient population may not reflect national sex distribution. If women are underrepresented in the training data, the model may perform worse in female patients due to differential inflammatory responses, body composition affecting drug distribution, or sex-specific lung mechanics. Mitigation: Report and ensure balanced sex representation; conduct stratified performance analysis.

**4. Sedation Practice Variation**

Deep sedation suppresses cough reflex and may alter secretion clearance patterns. If sedation depth varies systematically by patient characteristics (e.g., age, frailty), this unmeasured confounder could introduce bias. Mitigation: Include sedation depth scores (RASS, Richmond) as predictors.

**5. Temporal Bias and Pandemic-Era Practice Changes**

All 19 positive outcomes occurred post-2020, coinciding with COVID-19 pandemic practice changes including increased prone positioning, different antimicrobial protocols, and altered PCT measurement frequency. The model may not generalize to pre-pandemic or post-pandemic practice patterns. Mitigation: Validate on contemporary cohorts and update periodically.

##### Proposed Fairness Metrics for Validation

Multi-site validation should report:

• Equalized odds: sensitivity and specificity stratified by protected attributes

• Demographic parity: positive prediction rate stratified by protected attributes

• Calibration-in-the-large: calibration slope and intercept for each subgroup

• Subgroup sample sizes with confidence interval widths

##### Socioeconomic Context

The Dudley population includes significant socioeconomic deprivation (35% in IMD deciles 1-3), which may affect health-seeking behaviour, baseline health status, and healthcare access. While deprivation itself was not a model input, it may correlate with unmeasured confounders (smoking, nutrition, housing quality) that influence VAP risk. Generalizability to affluent catchment areas is uncertain.

#### Section S5: Complete Limitations with Mitigation Strategies

This section provides the complete list of study limitations with detailed discussion and proposed mitigation strategies for future work.

##### Limitation 1: Small Positive Sample Size

**Description:**

The final dataset comprised only 19 positive outcomes (0.82% prevalence), with 5 positive cases in the test set. This yields wide confidence intervals (sensitivity: 33.3-100.0%, PPV: 0.5-4.6%) reflecting fundamental statistical uncertainty when drawing inferences from five events.

**Impact:**

Limited precision in performance estimates. Point estimates (AUROC 0.851, sensitivity 80%) are uncertain. Test set results should be interpreted as preliminary evidence requiring validation.

**Mitigation:**

Multi-site pooled validation targeting ≥100 positive outcomes for narrow confidence intervals and robust performance characterization.

##### Limitation 2: Composite Surrogate Outcome

**Description:**

The outcome (≥5% P:F decline AND ≥0.5 ng/mL PCT rise) is not a validated VAP diagnostic label. Non-infective causes of respiratory deterioration (atelectasis, pulmonary oedema, ARDS progression, pulmonary embolism) could satisfy these criteria.

**Impact:**

Unknown sensitivity and specificity against gold-standard VAP diagnosis. The model predicts physiological deterioration that may correlate with, but not definitively indicate, VAP.

**Mitigation:**

Validate against: (1) microbiological VAP diagnosis (positive BAL/mini-BAL), (2) clinical VAP diagnosis (CPIS ≥6), (3) antibiotic initiation for suspected VAP, (4) radiologically-confirmed pneumonia.

##### Limitation 3: Low Retention Rate (12.8%)

**Description:**

Only 2,134 of 33,208 candidate blocks (6.4%) met all inclusion criteria, primarily driven by PCT availability (post-2020 only).

**Impact:**

Temporal bias with all 18 positive outcomes in the recent cohort. Performance on historical data unknown.

**Mitigation:**

Validate on contemporary cohorts with routine daily PCT measurement.

##### Limitation 4: Missing Treatment Data

**Description:**

Antibiotic therapy, sedation, neuromuscular blockade, and prone positioning not extracted.

**Impact:**

Unmeasured confounding. Antibiotics could suppress PCT independently of VAP resolution.

**Mitigation:**

Include treatment variables in future models; conduct sensitivity analysis.

##### Limitation 5: No Demographics or Severity Scores

**Description:**

No patient-level age, sex, ethnicity, APACHE II, or SOFA scores.

**Impact:**

Cannot assess fairness, case-mix adjustment, or subgroup performance.

**Mitigation:**

Mandatory demographic and severity data collection for validation.

##### Limitation 6: Single-Centre Threshold

**Mitigation:**

Develop site-specific threshold recalibration methodology.

##### Limitation 7: No Prospective Validation

**Mitigation:**

Prospective silent deployment with real-time performance monitoring.
